# Healthcare workers’ SARS-CoV-2 infection rates during the second wave of the pandemic: prospective follow-up study

**DOI:** 10.1101/2021.11.17.21266459

**Authors:** Anne Mette Würtz, Martin B. Kinnerup, Kirsten Pugdahl, Vivi Schlünssen, Jesper Medom Vestergaard, Kent Nielsen, Christine Cramer, Jens Peter Bonde, Karin Biering, Ole Carstensen, Karoline Kærgaard Hansen, Annett Dalbøge, Esben Meulengracht Flachs, Mette Lausten Hansen, Ane Marie Thulstrup, Else Toft Würtz, Mona Kjærsgaard, Mette Wulf Christensen, Henrik Albert Kolstad

## Abstract

**Objectives:** To assess if healthcare workers during the second wave of the coronavirus disease 2019 (COVID-19) pandemic had increased severe acute respiratory syndrome coronavirus 2 (SARS-CoV-2) infection rates following close contact with patients, co-workers and persons outside work with COVID-19.

**Methods:** A prospective cohort study of 5985 healthcare workers from Denmark were followed November 2020 to April 2021 and provided day-by-day information on COVID-19 contacts. SARS-CoV-2 infection was defined by the first positive polymerase chain reaction (PCR) test ever.

**Results:** 159 positive and 35 996 negative PCR tests were recorded during 514 165 person-days. The SARS-CoV-2 infection rate following close contact with COVID-19 patients 3-7 days earlier was 153.7 per 100,000 person-days corresponding with an incidence rate ratio (IRR) of 3.17 (40 cases, 95% CI 2.15 - 4.66) compared with no close contact. IRRs following close contact with co-workers and persons outside work with COVID-19 were 2.54 (10 cases, 95% CI 1.30 - 4.96) and 17.79 (35 cases, 95% CI 12.05 - 26.28). The estimates for close contact with COVID-19 patients, co-workers or persons outside work were mutually adjusted.

**Conclusions:** Despite strong focus on preventive measures during the second wave of the pandemic, healthcare workers were still at increased risk of SARS-CoV-2 infection when in close contact with patients with COVID-19. Among all health care workers, the numbers affected due to close patient contact were comparable to the numbers affected following COVID-19 contact outside work.

## Introduction

The first wave of the SARS-CoV-2 pandemic was globally characterised by widespread lack of personal protective equipment (PPE), confusing PPE guidelines and lack of SARS-CoV-2 testing and contact tracing (1). Healthcare workers were at highly increased risk of COVID-19 (2-4). March - April 2020, front-line healthcare workers in UK and USA reporting adequate PPE use when in direct contact with COVID-19 patients showed a five-fold increased self-reported positive polymerase chain reaction (PCR) testing rate for SARS-CoV-2 of 553 per 100,000 (3). Increased SARS-CoV-2 seropositivity was reported among healthcare workers in close contact with patients (5-7), co-workers (5, 6), household members and other persons outside work with COVID-19 (5, 6, 8-11), but not consistently (9-11).

A considerable increase in preventive measures was initiated in multiple countries including Denmark (12), and it was expected that the pandemic afflicting so many healthcare workers was brought under control during the second wave. We studied healthcare workers’ SARS-CoV-2 infection and COVID-19 symptom rates during the second wave of the pandemic following close contact with patients, co-workers and persons outside work with COVID-19 compared with no such contacts.

## Methods

### Study design and population

This is a dynamic prospective follow-up study with day-by-day self-reported information on COVID-19 contacts. Outcome is incident SARS-CoV-2 infection. Person-day is the unit of analysis. The study population is healthcare workers and technical, administrative and other staff of the Central Denmark Region (hereafter named healthcare workers).

### General surveillance and infection control recommendations

PCR testing for SARS-CoV-2 was freely accessible at no cost for all Danish citizens independent of symptoms. All hospital workers with any patient contact were urged to be PCR tested bi-weekly until January 26, 2021, thereafter weekly. PCR test results were provided on average 24-36 hours after sample collection. SARS-CoV-2 infection rates in the second wave of the pandemic peaked in Denmark December 16, 2020, with 4387 PCR verified cases in a population of 5,771,877 citizens.

All healthcare workers were instructed to follow general guidelines for infection control and wear surgical masks in all indoor areas with public or patient access and maintain physical distance to other persons whenever possible. All workers with non-critical functions were sent home December 11, 2021, and for the remaining study period.

During care for patients diagnosed with or under suspicion of COVID-19, all staff were instructed to wear a fluid repellent disposable gown with long sleeves, disposable medical gloves, surgical mask and protective glasses or visor. Moreover, during procedures with risk of aerosol generation (e.g. high flow oxygen therapy) the surgical mask should be replaced by a filtering face piece 2 or 3 (FFP2, FFP3) respirator. There was sufficient supply of PPE during the study period.

Following close contact with persons diagnosed with COVID-19 without prescribed PPE for 15 minutes or more, any citizen had to go into self-isolation and be PCR tested at day four and six. Self-isolation could be cancelled following two negative tests or, in case of a positive test, 48 hours after symptom cessation or seven days after the positive test if asymptomatic. Detailed infection control for COVID-19 for employees of the Central Denmark Region during the COVID-19 pandemic can be found in the supplemental material.

### Exposure assessment of COVID-19 contacts

Each day during follow-up at 3:30 pm, study participants received a text message linking to a questionnaire. They were asked to report any incident of close contact within a one-meter distance with patients and persons outside work with COVID-19 during the current and the previous 1-2 and 3-4 days. Participants were also asked to report incidents of close contact with co-workers with COVID-19 during the previous 1-2 and 3-4 days, but not the current day, because co-workers with known COVID-19 would not be present at work.

For each day in the follow-up period, we assigned exposure individually based on participants’ own reporting of close COVID-19 contacts in a 5-day time window starting seven days and ending three days before each day of follow-up to account for the expected latent period (13). Days were classified with close contact with COVID-19 patients if the participant reported such contact at least once during the 5-day time window. Otherwise, days were classified with no close contact with COVID-19 patients if participants reported this for three or more days during the 5-day window. Days not fulfilling these criteria were classified with unknown close contact with COVID-19 patients. Days of follow-up were classified in the same way following close contact with co-workers and persons outside work.

### SARS-CoV-2 infection, vaccination and COVID-19 symptoms

The main outcome measure was incident SARS-CoV-2 infection defined as the first positive PCR test ever recorded in a regional register with complete coverage of all tests conducted in the population since February 27, 2020. A regional register also provided information about all COVID-19 vaccinations since December 27, 2020. The secondary outcome measure was first report of loss of taste and smell as asked for in the daily questionnaire.

### Population characteristics

Information on age, sex, occupation and department of employment was obtained from the personnel records of the Central Denmark Region. Information on smoking, BMI, airways disease (chronic obstructive pulmonary disease, asthma, rhinitis) was reported by the participants at baseline. Non-compliance with PPE guidelines was reported in the daily questionnaire.

### Statistical analyses

Study participants were followed day-by-day from seven days after the first daily questionnaire response, November 25, 2020, at the earliest, until first positive test for SARS-CoV-2, seven days after full vaccination (14) or April 30, 2021. Each day of follow-up was classified according to close contact (yes, no) with patients, co-workers and persons outside work with COVID-19 according to the previously defined criteria. Participants may have experienced all contact forms several times during follow-up and thus move in and out of exposures.

We used generalised linear models with log-link assuming a Poisson distribution with person-days as offset representing the time at risk to derive incidence rate ratios (IRRs) with 95% confidence intervals (CIs) for SARS-CoV-2 infection. Adjusted IRRs were mutually adjusted for the other types of COVID-19 contact, sex, age (continuous) and month (6 categories, November 2020-April 2021) as decided a priori. We furthermore adjusted for number of PCR tests made before, during and after the 5-day exposure window (≥ 8, 3-7 and 1-2 days previously). However, this only affected IRR estimates marginally, and in the final models, we included the cumulative number of earlier PCR tests as a continuous variable. This and all other variables were treated as time-varying day-by-day.

We excluded person-days with missing information on close contact with patients, co-workers and persons outside work diagnosed with COVID-19. We abstained from imputing the missing values. This was because a high fraction of participants worked part time or irregular shifts with at least two days off work with no close contact with patients or co-workers at unpredictable days during a given week. Information on the covariates of the adjusted models were complete.

Analyses of loss of taste and smell followed a similar setup as SARS-CoV-2 infection, but we did not censor subjects when testing PCR positive for SARS-CoV-2 and did not include number of earlier PCR tests in the adjusted models.

In a sensitivity analysis of possible differential recall of close COVID-19 contacts, we excluded contact information obtained after a given day of follow-up (i.e. based on questionnaire reports for the previous 1-2 and 3-4 days), when PCR test results were available for the participants. This excluded information on close contact with co-workers with COVID-19 because this was only reported for the previous 1-2 and 3-4 days.

## Results

A total of 26,089 healthcare workers were invited to the study November 17, 2020. After excluding 724 who were positive for SARS-CoV-2 before start of follow-up, 25,365 healthcare workers (3,253671 person-days) were candidates for inclusion and 6337 (753,607 person-days) participated (Table 1). After excluding person-days with missing information on close contact with patients, co-workers or persons outside work with COVID-19, the study population included 5985 healthcare workers providing 514,165 person-days at risk. The daily testing rates were 5.5% for the invited population and 7.1% for the study population. Altogether, 448,748 daily questionnaire responses were collected from the study population during follow-up, corresponding with an 87.3% coverage. SARS-CoV-2 infection rates in the invited population and the study population were 28.6 and 30.9 per 100,000 person-days.

**Table 1.**
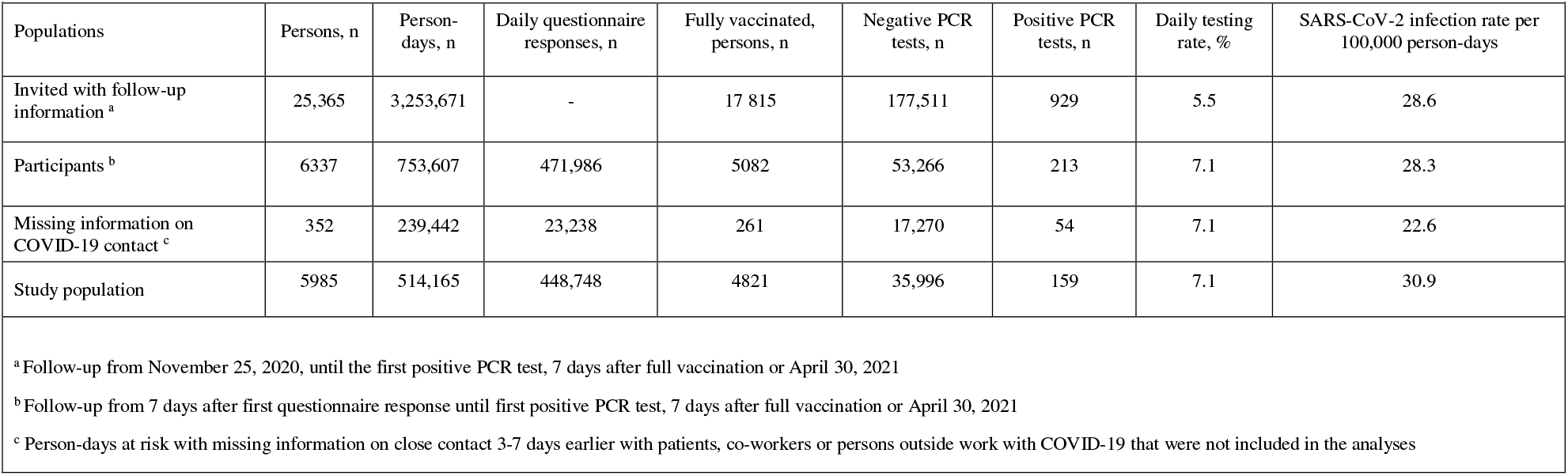
Study profile

Table 2 presents characteristics (person-days) of the invited population and the study population by COVID-19 contacts 3-7 days earlier. The study population included 88.6% women and the mean age was 48.0 years compared with 83% women and a mean age of 43.6 years for the invited population. More study participants compared with the invited healthcare workers had been PCR tested earlier. Only minor occupation and department differences between the invited and the participating populations were seen.

**Table 2.** Population characteristics (person-days) according to participation status and contact 3-7 days earlier with patients, co-workers and persons outside work with COVID-19

|  |  |  |  |  |  |  |  |
| --- | --- | --- | --- | --- | --- | --- | --- |
|  |  | COVID-19 contact among participants |  |  |  |  |  |
|  | Invited population | Patients |  | Co-workers |  | Persons outside work |  |
| Characteristics | n = 3,253,671 | No (n = 488 147) | Yes (n = 26 018) | No (n = 510 012) | Yes (n = 4 153) | No (n = 509 358) | Yes (n = 4 807) |
| Women n (%) | 2 708,710 (83) | 435,986 (89) | 22,650 (87) | 455,003 (89) | 3633 (87) | 454,360 (89) | 4276 (89) |
| Age, mean (SD) | 43.4 (12.1) | 49.5 (10.3) | 47.3 (11.1) | 49.4 (10.4) | 49.2 (10.6) | 49.4 (10.4) | 48.4 (10.5) |
| COVID-19 contact, n (%) |  |  |  |  |  |  |  |
| Patients | - | - | - | 24,658 (5) | 1360 (33) | 25,504 (5) | 514 (11) |
| Co-workers | - | 2793 (1) | 1360 (5) | - | - | 4039 (1) | 114 (2) |
| Persons outside work | - | 4293 (1) | 514 (2) | 4693 (1) | 114 (3) | - | - |
| Months, n (%) |  |  |  |  |  |  |  |
| November | 760,392 (23) | 15,286 (3) | 922 (4) | 16,039 (3) | 169 (4) | 16,005 (3) | 203 (4) |
| December | 571,757 (18) | 129,414 (27) | 9527 (37) | 136,712 (27) | 2229 (54) | 135,885 (27) | 3056 (64) |
| January | 535,177 (16) | 126,987 (26) | 11,459 (44) | 136,899 (27) | 1547 (37) | 137,437 (27) | 1009 (21) |
| February | 456,640 (14) | 85,888 (18) | 2507 (10) | 88,306 (17) | 89 (2) | 88,222 (17) | 173 (4) |
| March | 152,046 (5) | 73,007 (15) | 913 (4) | 73,861 (14) | 59 (1) | 73,759 (14) | 161 (3) |
| April | 777,659 (24) | 57,565 (12) | 690 (3) | 58,195 (11) | 60 (1) | 58,050 (11) | 205 (4) |
| PCR tests, 1-2 days earlier, n (%) |  |  |  |  |  |  |  |
| 0 | 2,909,178 (89) | 421,861 (86) | 21,117 (81) | 440,408 (86) | 2570 (62) | 439,936 (86) | 3042 (63) |
| 1-2 | 343,106 (11) | 66,108 (14) | 4808 (19) | 69,362 (14) | 1554 (38) | 69,176 (14) | 1740 (37) |
| PCR tests, 3 to 7 days earlier, n (%) |  |  |  |  |  |  |  |
| 0 | 2,460,903 (76) | 334,339 (68) | 15,461 (59) | 347,771 (68) | 2029 (49) | 347,449 (68) | 2351 (49) |
| 1 | 753,731 (23) | 147,119 (30) | 9269 (36) | 154,621 (30) | 1767 (43) | 154,484 (30) | 1904 (40) |
| ≥2 | 39,037 (1) | 6,689 (1) | 1288 (5) | 7620 (1) | 357 (9) | 7425 (1) | 552 (11) |
| PCR tests, ≥ 8 days earlier, n (%) |  |  |  |  |  |  |  |
| 0 | 647,158 (20) | 53,070 (11) | 1921 (7) | 54,586 (11) | 405 (10) | 54,415 (11) | 576 (12) |
| 1 – 4 | 1,346,111 (41) | 215,792 (44) | 11,404 (44) | 224,818 (44) | 2378 (57) | 224,339 (44) | 2857 (59) |
| 5 – 9 | 861,939 (26) | 141,699 (29) | 9720 (37) | 150,303 (29) | 1116 (27) | 150,374 (30) | 1045 (22) |
| ≥ 10 | 398,463 (12) | 77,586 (16) | 2973 (11) | 80,305 (16) | 254 (6) | 80,230 (16) | 329 (7) |
| Occupation, n (%) |  |  |  |  |  |  |  |
| Nursing staff <sup>a</sup> | 1,264,494 (39) | 180,659 (37) | 13,532 (52) | 192,065 (38) | 2126 (51) | 192,077 (38) | 2114 (44) |
| Medical doctors | 451,218 (14) | 43,574 (9) | 3092 (12) | 46,275 (9) | 391 (9) | 46,184 (9) | 482 (10) |
| Biomedical Laboratory | 156,073 (5) | 36,657 (8) | 3398 (13) | 39,788 (8) | 267 (6) | 39,769 (8) | 286 (6) |
| Medical secretaries | 277,011 (9) | 61,854 (13) | 864 (3) | 62,357 (12) | 361 (9) | 62,091 (12) | 627 (13) |
| Other | 1,094,735 (34) | 165,260 (34) | 5127 (20) | 169,379 (33) | 1008 (24) | 169,089 (33) | 1298 (27) |
| Missing | 10,140 (0) | 143 (0) | 5 (0) | 148 (0) | 0 (0) | 148 (0) | 0 (0) |
| Department, n (%) |  |  |  |  |  |  |  |
| Emergency | 120,912 (4) | 9,413 (2) | 2509 (10) | 11,602 (2) | 320 (8) | 11,686 (2) | 236 (5) |
| Medicine <sup>b</sup> | 776,923 (24) | 123,542 (25) | 6557 (25) | 128,932 (25) | 1167 (28) | 128,827 (25) | 1272 (26) |
| Surgery <sup>c</sup> | 603,097 (19) | 86,542 (18) | 2897 (11) | 88,877 (17) | 562 (14) | 88,581 (17) | 858 (18) |
| Biochemistry | 184,158 (6) | 38,959 (8) | 3273 (13) | 42,018 (8) | 214 (5) | 41,942 (8) | 290 (6) |
| Service <sup>d</sup> | 109,284 (3) | 6,509 (1) | 651 (3) | 7104 (1) | 56 (1) | 7,137 (1) | 23 (0) |
| Anaesthesiology | 123,557 (4) | 16,121 (3) | 3325 (13) | 19,169 (4) | 277 (7) | 19,246 (4) | 200 (4) |
| Radiology and Nuclear Medicine | 134,114 (4) | 21,574 (4) | 1500 (6) | 22,826 (4) | 248 (6) | 22,821 (4) | 253 (5) |
| Psychiatry | 427,205 (13) | 61,721 (13) | 1266 (5) | 62,474 (12) | 513 (12) | 62,343 (12) | 644 (13) |
| Departments with less frequent patient contact <sup>e</sup> | 455,684 (14) | 92,302 (19) | 2483 (10) | 94,281 (18) | 504 (12) | 94,051 (18) | 734 (15) |
| Other <sup>f</sup> | 308,597 (9) | 31,321 (6) | 552 (6) | 32,581 (6) | 292 (7) | 32,576 (6) | 297 (6) |
| Missing | 10,140 (0) | 143 (0) | 5 (0) | 148 (0) | (0) | 148 (0) | 0 (0) |
| Smoking, n (%) |  |  |  |  |  |  |  |
| Current smoker | - | 26,498 (5) | 1951 (7) | 28,222 (6) | 227 (5) | 28,150 (6) | 299 (6) |
| Previous smoker | - | 142,411 (29) | 7660 (29) | 148,682 (29) | 1389 (33) | 148,549 (29) | 1522 (32) |
| Never smoker | - | 315,544 (65) | 16,251 (62) | 329,287 (65) | 2508 (60) | 328,822 (65) | 2973 (62) |
| Missing | - | 3694 (1) | 156 (1) | 3821 (1) | 29 (1) | 3837 (1) | 13 (0) |
| BMI (kg/m<sup>2</sup>),n (%) |  |  |  |  |  |  |  |
| <20 | - | 32,317 (7) | 1710 (7) | 33,764 (7) | 263 (6) | 33,715 (7) | 312 (6) |
| 20-24 | - | 228,612 (47) | 11,608 (45) | 238,506 (47) | 1714 (41) | 237,771 (47) | 2449 (51) |
| 25-29 | - | 146,236 (30) | 7777 (30) | 152,586 (30) | 1427 (34) | 152,662 (30) | 1351 (28) |
| ≥30 | - | 77,116 (16) | 4734 (18) | 81,143 (16) | 707 (17) | 81,168 (16) | 682 (14) |
| Missing | - | 3,866 (1) | 189 (1) | 4013 (1) | 42 (1) | 4042 (1) | 13 (0) |
| Lung disease |  |  |  |  |  |  |  |
| Hay fever | - | 100,096 (21) | 4827 (19) | 104,107 (20) | 816 (20) | 103,924 (20) | 999 (21) |
| Asthma | - | 34,659 (7) | 1668 (6) | 36,036 (7) | 291 (7) | 35,900 (7) | 427 (9) |
| COPD <sup>g</sup> | - | 3091 (1) | 188 (1) | 3232 (1) | 47 (1) | 3231 (1) | 48 (1) |
| <sup>a</sup> Nurses, social- and healthcare assistants and radiographers<br><sup>b</sup> Internal medicine, paediatrics, oncology and neurology<br><sup>c</sup> All surgical departments, including: obstetrics and gynaecology; otorhinolaryngology, head and neck surgery; and ophthalmology<br><sup>d</sup> Cleaning services; hospital porters; clothing and waste management; depot and archive; telephone switchboard; and guidance for patients, relatives and staff<br><sup>e</sup> Occupational and social medicine; physio- and occupational therapy; administration; department of technical services; and kitchen<br><sup>f</sup> Administrative, technical and pedagogical staff<br><sup>g</sup> COPD Chronic obstructive pulmonary disease |  |  |  |  |  |  |  |

Participants with one type of close COVID-19 contact more often also reported the other types of close COVID-19 contact. All types of COVID-19 contact were associated with more frequent PCR testing, especially during the previous 1-2 days. More nurses had close contact with patients and co-workers with COVID-19 than other occupations. Only small differences were seen for department, smoking status, BMI and lung diseases.

Forty participants tested positive for SARS-CoV-2 after having close contact with COVID-19 patients 3-7 days earlier (Table 3). This gave an infection rate of 153.7 per 100,000 person-days and an adjusted IRR of 3.17 (95% CI 2.15 - 4.66) when compared with no close contact with COVID-19 patients. Ten and 35 participants had close contact with co-workers and persons outside work with COVID-19, respectively, giving infection rates of 240.8 and 728.1 per 100,000 person-days and adjusted IRRs of 2.54 (95% CI 1.30 - 4.96) and 17.79 (95% CI 12.05 - 26.28).

**Table 3.** Close contact 3-7 days earlier with patients, co-workers and persons outside work with COVID-19 and incidence rate ratios of SARS-CoV-2

|  |  |  |  |  |  |  |  |
| --- | --- | --- | --- | --- | --- | --- | --- |
| Contact with persons with COVID-19 | Person-days | First positive SARS-CoV-2 PCR tests | Infection rate per 100,000 person-days | Incidence rate ratios (95% CI) |  |  |  |
|  |  |  |  | Model 1* | Model 2† | Model 3‡ | Model 4§ |
| Patients |  |  |  |  |  |  |  |
| No contact | 488,147 | 119 | 24.4 | Reference | Reference | Reference | Reference |
| Contact | 26,018 | 40 | 153.7 | 6.31 (4.41 - 9.02) | 4.62 (3.21 - 6.65) | 3.72 (2.55 - 5.44) | 3.17 (2.15 - 4.66) |
| Co-workers |  |  |  |  |  |  |  |
| No contact | 510,012 | 149 | 29.2 | Reference | Reference | Reference | Reference |
| Contact | 4153 | 10 | 240.8 | 8.24 (4.34 - 15.63) | 5.44 (2.86 - 10.35) | 2.68 (1.37 - 5.24) | 2.54 (1.30 - 4.96) |
| Persons outside work |  |  |  |  |  |  |  |
| No contact | 509,358 | 124 | 24.3 | Reference | Reference | Reference | Reference |
| Contact | 4807 | 35 | 728.1 | 29.91 (20.55 - 43.52) | 21.75 (14.75 - 32.06) | 18.87 (12.78 - 27.88) | 17.79 (12.05 - 26.28) |
| *Crude model |  |  |  |  |  |  |  |
| †Adjusted for age (continuous), sex and month (6 categories, November 2020-April 2021) |  |  |  |  |  |  |  |
| ‡As model 2 and additionally adjusted for the other types of COVID-19 contact |  |  |  |  |  |  |  |
| § As model 3 and additionally adjusted for number of previous PCR tests. |  |  |  |  |  |  |  |

A total of 24 participants with incident loss of taste and smell had experienced close contact with COVID-19 patients (Table 4). This corresponded with an incidence rate of 41.4 per 100,000 person-days and an adjusted IRR of 1.48 (95% CI 0.95 - 2.29) (Table 4). Following close contact with co-workers and persons outside work with COVID-19, the adjusted IRRs of loss of taste and smell were 2.56 (95% CI 1.24 - 5.30) and 10.82 (95% CI 7.33 - 15.98). Among those reporting loss of taste and smell, 36% had an earlier positive PCR test.

**Table 4.** Close contact 3-7 days earlier with patients, co-workers and persons outside work with COVID-19 and incidence rate ratios of loss of taste and smell

|  |  |  |  |  |  |  |
| --- | --- | --- | --- | --- | --- | --- |
| Close contact with persons with COVID-19 | Person-days* | Incident loss of taste and smell | Incidence rate per 100,000 person-days | Incidence rate ratios (95% CI) |  |  |
|  |  |  |  | Model 1<sup>†</sup> | Model 2<sup>‡</sup> | Model 3<sup>§</sup> |
| Patients |  |  |  |  |  |  |
| No contact | 488,451 | 202 | 41.4 | Reference | Reference | Reference |
| Contact | 26,748 | 24 | 89.7 | 2.17 (1.42 to 3.31) | 1.70 (1.11 to 2.61) | 1.48 (0.95 to 2.29) |
| Co-workers |  |  |  |  |  |  |
| No contact | 511,010 | 218 | 42.7 | Reference | Reference | Reference |
| Contact | 4189 | 8 | 191.0 | 4.48 (2.21 to 9.07) | 3.20 (1.57 to 6.49) | 2.56 (1.24 to 5.30) |
| Persons outside work |  |  |  |  |  |  |
| No contact | 510,123 | 195 | 38.2 | Reference | Reference | Reference |
| Contact | 5076 | 31 | 610.7 | 15.98 (10.94 to 23.34) | 11.21 (7.60 to 16.54) | 10.82 (7.33 to 15.98) |
| *This population was slightly different from that of table 3 because of the different outcome<br><sup>†</sup>Crude model<br><sup>‡</sup>Adjusted for age (continuous), sex and month (6 categories, November 2020-April 2021)<br><sup>§</sup> As model 2 and additionally adjusted for the other types of COVID-19 contact |  |  |  |  |  |  |

The infection rate in the study population declined from January to April 2021, increased by number of PCR tests 3-7 days earlier and were higher for departments of medicine and among nurses compared with other departments and occupations. No clear infection rate patterns were seen for the other population characteristics (Supplementary table S1).

Participants reported an overall 2% non-compliance with PPE guidelines during 187,413 daily procedures. For respiratory procedures with potential for higher exposure levels, this percentage was 4.8% (Supplementary table S2).

Sensitivity analyses that only included COVID-19 contact information obtained before results of the PCR tests were available, showed an infection rate of 155.2 per 100,000 person-days and an adjusted IRR of 3.52 (95% 2.41 - 5.13) following close contact with COVID-19 patients (Supplementary table S3). The IRR following close contact with persons outside work with COVID-19 was 14.19 (95% CI 8.27 - 24.33). No results were available for close contact with co-workers with COVID-19 because this information was obtained after PCR test results were available for those tested.

## Discussion

### Principal findings

This prospective follow-up study was conducted from November 25, 2020 to April 30, 2021 during the second wave of the pandemic in Denmark. The SARS-CoV-2 infection rate following close contact with COVID-19 patients was 153.7 per 100,000 person-days. This corresponded with a three-fold increased adjusted IRR compared with no such contact. Close contact with persons outside work with COVID-19 showed an almost 18-fold increased infection rate. The absolute number of healthcare workers affected were similar following contact with patients and persons outside work with COVID-19. Contact with co-workers was also associated with an increased risk of SARS-CoV-2 infection. Comparable patterns of increased risks of loss of taste and smell were seen for all three types of COVID-19 contact. Participants reported high but not complete day-by-day compliance with PPE guidelines.

### Strengths and limitations

Strengths of this study are the prospective follow-up design with day-by-day information that allowed precise account for latent period and day-by-day change in exposure, the complete follow-up for PCR test results, and information on incident loss of taste and smell that is a signature of SARS-CoV-2 infection (15). During Spring 2020, persistent loss of taste and smell was in this population strongly associated with a positive PCR test for SARS-CoV-2 with an odds ratio of 57.16 (95% CI 16.71 - 195), corresponding with a specificity of 98% and a positive predictive value of 84% (16). Other strengths are the free access to PCR testing and the high testing rate. The decision to be PCR tested was therefore unlikely to be strongly associated with COVID-19 contact and result of the PCR test, and we regard collider bias a minor problem (17).

Participants were tested more often and showed a higher infection rate than the invited population. Otherwise, participants were comparable with the invited population and this neither suggests strong collider bias (17).

Participants reporting one type of close COVID-19 contact (patients, co-workers or persons outside work) more often experienced the other types of contact and the mutually adjusted IRR estimates were substantially reduced and are expected to provide the best estimates of the separate effects. Participants with close COVID-contacts had been PCR tested for SARS-CoV-2 infection more often than those with no such contacts. However, earlier PCR tests (all negative) should not be causally associated with SARS-CoV-2 infection as detected by a positive PCR test on a given day of follow-up but may be indicators of unobserved risk factors of SARS-CoV-2 infection that may confound associations. Our analyses indicated no such confounding.

COVID-19 contact information was partly obtained after the results of the PCR test results were available for the tested participants, which may have introduced recall bias and inflated results. However, sensitivity analyses relying only on contact information obtained before results of the PCR tests were available indicated no substantial recall bias. Knowledge of PCR test results as well as COVID-19 contact may, on the other hand, have inflated results for loss of taste and smell. Being classified with no close COVID-19 contact during the 5-day exposure window allowed missing information for two of the five days. Because there may have been COVID-19 contact during these days, this may have attenuated IRRs.

The low number of SARS-CoV-2 infected participants reduced the statistical strength and thus restricted the number of potential confounders taken into account.

### Comparisons with other studies

This study showed an overall SARS-CoV-2 infection rate of 30.9 per 100,000 person-days, which was below the self-reported positive PCR testing rates of 132 per 100,0000 person-days observed in a prospective cohort of frontline healthcare worker of the first wave by Nugyen et al (3). Our observed infection rate of 153.7 per 100,000 person-days following contact with COVID-19 patients was also lower than the 553 per 100,000 person-days reported by Nugyen et al. following such contact among healthcare workers reporting adequate PPE use (3). These findings are, however, not directly comparable with ours because of differences in population compositions and definitions of COVID-19 contact and SARS-CoV-2 infection.

Norwegian nurses and physicians showed a tree-fold increased odds of SARS-CoV-2 infection during the first wave (26 February – 17 July 2020) compared with the general working population (18). During the second wave (18 July – 18 December 2020), odds ratios were well below 1.5 for these two occupations (18).

We observed a SARS-CoV-2 infection rate of 728.1 per 100,000 person-days following close contact with persons outside work with COVID-19, which was half the average household infection rate of 1660 per 100,000 person-days reported for the first wave (8). This may partly reflect that we included any close contact with a person outside work with COVID-19 and not only household contacts that are expected to be closer and last longer.

### Concluding remarks

During the second wave of the pandemic, this healthcare worker population was at increased risk of SARS-CoV-2 infection when in close contact with COVID-19 patients. Among all healthcare workers, the absolute numbers affected were similar to the absolute numbers affected following COVID-19 contact outside work. Close contact with co-workers also entailed an increased risk. PPE was not in shortage, guidelines for proper PPE use and other infection control measures were implemented and compliance with required PPE was high but not complete. Vaccination will not eliminate risks (19). The current findings thus stress the need for increased focus on use of recommended PPE, correct donning, doffing and other procedures (20-22), training (23) and ventilation (24). The aim is to secure healthcare workers’ health and reduce transmission into the community (25) during ongoing and future waves of SARS-CoV-2 and other infections.

## Data Availability

No additional data available. For legal and ethical reasons, individual level patient data cannot be shared by the authors and are only accessible to authorised researchers after application to the Danish Health Data Authority.

## Acknowledgements

This research has received funding from Central Denmark Region (grant number RR 20200527) and Danish Working Environment Fund (grant number 20205100734). All authors declare no conflicts of interest.

