## Supplementary material for "Healthcare workers’ SARS-CoV-2 infection rates during the second wave of the pandemic: prospective follow-up study"

**COVID-19. Infection control precautions, regional guideline**

Urgent treatment and diagnostics must not be delayed due to any suspicion of COVID-19.

The number of persons in contact with the patient should be kept to a necessary minimum. Donning and removal of personal protective equipment (PPE) should be trained. For choice of PPE see Appendix 1: Choice of personal protective equipment for suspected or verified COVID-19

General precautions always apply. In addition, additional precautions corresponding to contact infection and droplet infection are used.

Schema 1 and Schema 2 below describe the additional infection control measures for suspected or confirmed COVID-19. Schema 3 describes tracing of close contacts as well as containment and outbreak management in case of detection of SARS-CoV-2 RNA in staff and patients.

**Schema 1: Inpatient hospital ward**

| | |
|---|---|
| *Route of transmission* | Droplet transmission and direct and indirect contact transmission. |
| *Additional precautions apply to:* | **Patients with symptoms of COVID-19 (suspected or verified COVID-19)**<br>• Suspicion of COVID-19 should arise in the spectrum from mild symptoms of upper and lower respiratory tract infection to severe lower respiratory infection.<br>• Suspicion of COVID-19 should arise **also** in vaccinated patients<br>• Suspicion of COVID-19 should arise **also** in patients previously tested positive for SARS-CoV-2 RNA.<br><br>The additional precautions apply for patients with symptoms until the criteria in the section "Duration of isolation" are fulfilled.<br><br>**Patients without symptoms of COVID-19, who have tested PCR positive for SARS-CoV-2 RNA:**<br>• Patients who have never previously had a positive SARS-CoV-2 RNA test until 7 days after the positive test, provided they are still without symptoms.<br>• Patients, who are past a verified COVID-19 infection (>48 hours after end of symptoms or after expert opinion) or previously tested SARS-CoV-2 RNA positive, if more than 12 weeks have passed since end of symptoms (asymptomatic) or 12 weeks after first positive SARS-CoV-2 RNA test (asymptomatic)<br>•<br>**Patients who are close contacts** to a person tested positive for SARS-CoV-2 RNA:<br><br>• <u>If the exposure has ended</u> additional precautions apply until showing a negative test for SARS-CoV-2 RNA taken minimum 6 days after last exposure. |

| | |
|---|---|
|  | • <u>If the exposure continues</u> (e.g. for a parent taking care of a child tested positive for SARS-CoV-2 RNA) additional precautions apply until showing a negative test for SARS-CoV-2 RNA taken:<br> • at least 48 hours after the positive contact (the child) is symptom free or<br> • at least 7 days after the first positive test, if the positive contact (the child) is asymptomatic.<br><br>**NB:** Patients without symptoms of COVID-19, with a missing test result of the SARS-CoV-2 RNA test are not included in the additional precautions in this guideline. Please refer to "General infection control precautions – during an epidemic, regional guideline". |
| *Diagnostics of SARS-CoV-2* | See Department of Clinical Microbiology's direction for requisition "1.3. Corona virus (SARS-CoV-2 causing COVID-19) RNA – detection by PCR". |
| *Isolation in a single room – suspected or verified COVID-19* | Use a single room with a separate bathroom for patients with confirmed or suspected SARS-CoV-2 infection. The door should be marked with an isolation sign.<br><br>Isolation behind a room divider may be used for shorter stays, e.g. at recovery rooms. Isolation behind a room divider requires space and clear marking around the isolated patient, and adherence to general and supplementing control guidelines. The local infection control unit can be contacted for advice.<br><br>Room division between more isolated patients can if necessary be used in units with patients awaiting the COVID-19 diagnosis. These should be established in collaboration with the local infection prevention and control unit (IPCU).<br><br>Instruction for inpatients: "Patient instruction_isolation COVID-19.<br><br>Instruction for discharged patient: "For you who have been tested positive for new corona virus.<br><br>Isolation sign for the door of the room:<br>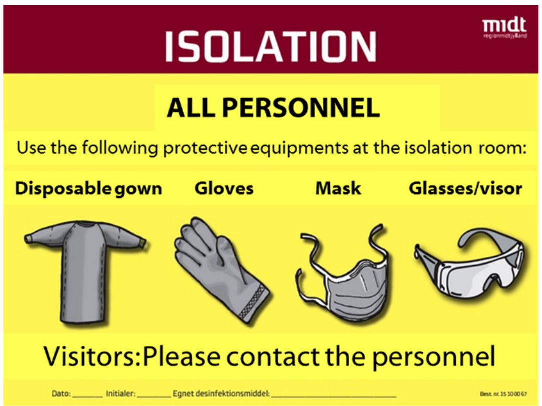<br>If aerosol generating procedures are performed in the room, the above sign should also be used for the door. |
| *Cohort isolation – patient with verified COVID-19* | Cohort isolation of patients with verified SARS-CoV-2 infection can be done if it is not possible to isolate the patients in single room.<br><br>Cohort isolation gives rise to special infection control challenges and should be done in collaboration with the local IPCU. |
| *Choice of personal* | For choice of personal protective equipment, see appendix:<br>"Choice of personal protective equipment for suspected or verified COVID-19" |

| | |
|---|---|
| *protective equipment for COVID-19* | The individual work task must be risk assessed with respect to duration and exposure for droplets, aerosols or moist generating procedures. If the risk of moist or contamination is high, a liquid tight gown and a full-face shield in addition to the face mask are chosen.<br><br>**Special for cohort isolation:** Disposable gowns can be used for more patients if changed between patients after exposure to contamination (e.g. person lifting or other direct contact with patients/equipment or aerosols).<br><br>To reduce the risk of contamination of the disposable gown:<br>• Protect the front by a plastic apron<br>• Protect the sleeves by oversleeves for close caring contacts.<br><br>Plastic apron and oversleeves are changed between patients.<br><br>**Special for cohort isolation of COVID-19 patients in intensive care:**<br><br>Intensive care involves several high-risk procedures for aerosol generation and the personnel are indicated to use a respirator. As the personnel by cohort isolation often goes from one patient to another, the indication for which mask to use can be difficult to fulfil. Also too many shifts between different masks are inexpedient.<br><br>It is thus recommended in "Note of aerosol generating procedures in the airways of patients with suspected or verified COVID-19" from the National Serum Institute, that all health personnel staying within 2 meters from cohort-isolated patients in intensive care with suspected or verified COVID-19 wear a FFP2/FFP3 respirator and eye protection.<br><br>**Special for testing units,** see appendices:<br><br>"personal protective equipment for testing units_throat swap by suspected COVID-19"<br>"Testing units – donning of personal protective equipment"<br>"Testing units – removal of personal protective equipment"<br><br>**Special for FFP2/3 respirators with valve**<br><br>Respirators with valve should be used <u>only</u> for patients with verified COVID-19, as the exhaled air is not filtered and the patient is not protected from infection from the personnel. By aerosol generating procedures in patients with suspected, but still unverified COVID-19, a FFP2/3 respirator without valve should be used. |
| *personal protective equipment – general principles* | For all work outside the isolation room, the general infection control precautions apply.<br><br>Mask and eye protection: Can be worn continuously by the personnel between patients, who are in isolation due to suspected or verified COVID-19. Mask and eye protection should not be touched, and should always be changed if visibly contaminated or moist. If the mask is removed, it should be disposed. If visor or protective googles are removed, they should be disposed or disinfected – see appendix "Range of personal protective equipment including disinfection guidance for COVID-19".<br><br>Regarding working environment in relation to personal protective equipment, se "Working environment and COVID-19/personal protective equipment.<br><br>The disposable gown should as a rule be changed between patients – see, however, special rules for cohort isolation and testing facilities. |

| | |
|---|---|
| | Gloves are changed between patients, and when changing from an unclean to a clean procedure in the same patient.<br><br>**Removal of personal protective equipment**<br><br>1. Gloves are removed followed by hand disinfection<br>2. Fluid repellent disposable gown with long sleeves are removed followed by hand disinfection<br>3. Visor/protective glasses are removed followed by hand disinfection<br>4. Masks are removed followed by hand disinfection<br><br>"Removal of personal protective equipment – FOR PRINT"<br><br>See in addition appendices:<br><br>"Donning and removal of disposable gown and gloves, Regional infection control guidelines"<br>"Donning and removal, Mask and visor with tie-strings, Regional infection control guidelines"<br>"Donning and removal. Respirator and protective goggles, regional infection control guidelines" |
| *Hand hygiene* | Hand disinfection are performed before leaving the isolation room.<br><br>Change of gloves and hand disinfection should be done between clean and unclean procedures, and for example before touching clean equipment in cupboards.<br><br>The patient is instructed in correct hand hygiene and assisted if necessary.<br><br>A sign regarding hand hygiene is placed on the inside of the isolation room door or at the indication of the cohort.<br><br>See "Hand disinfection" (for print). |
| *Visitors* | Visitors are instructed in hand hygiene and should use the same personal protective equipment as the personnel.<br><br>Personnel should help visitors with donning and removal of personal protective equipment.<br><br>Individuals in COVID-l9 related self-isolation may under special circumstances be allowed to visit the hospital. Contact the local IPCU for advice. |
| *Disinfectant* | Ethanol 70-85 % v/v (volume/volume percentage). |
| *Information* | Collaborating departments/personnel (medical laboratory technicians, service staff, radiographs etc.) should be informed about the additional precautions. |
| *Transportation in-house* | Before transportation:<br><br>• The patient should wear clean clothes and bed linen<br>• The patient should disinfect hands before leaving the room<br>• Any wounds should be covered by a tight, clean, and dry bandage<br>• Just before the transportation cleaned guardrails, headboards, and elevation control devices are disinfected.<br>• The patient should wear a type I or II face mask during the transport. If needed to do tasks in relation to the patient during the transport, the personnel should use the appropriate personal protective equipment. |

| | |
|---|---|
| | **Special for patient transport with ongoing procedures with high-risk of aerosol generation (e.g. high-flow oxygen therapy):**<br><br>• Choose as far as possible the least trafficked time for transportation and the least trafficked route.<br>• All personnel within 2-meter distance from the patient's airways should use gloves, long-sleeve disposable gown, FFP2 or FFP3 respirators, and protective goggles or visor.<br>• The transport should be accompanied by a least one clean assistant to handle necessary control panels, door handles and openers, and to assure that persons outside the transport team are keeping at least 2 meters distance.<br>• Immediately after the transport has passed, all contact point where the transport has stopped are cleaned and disinfected in a radius of 2 meters from the patient (e.g. in and outside the elevator). |
| *Transportation off-site* | The patient should wear a type I or II face mask during the transport.<br><br>When ordering an ambulance, ambulance aircraft or similar, inform of the additional infection hygiene measures that must be used for transfers or treatment tasks. |
| *Utensils* | Utensils are brought straight to the washer disinfector. Personal protective equipment (gloves, disposable gown, and sometimes face mask and eye protection) are removed before leaving the room or the cohort. Use new gloves and possibly a disposable apron when transporting the utensils to the wash room.<br><br>Utensils are heat disinfected in a washer disinfector. May be washed/disinfected with other utensils.<br><br>Toys: If toys belonging to the department are reused, it must be cleaned and disinfected as described above.<br><br>Apply to the general guidelines for cleaning/disinfection. More can be read here: "e-Dok Disinfection. Infection control precautions, regional guideline".<br><br>For reuse of protective goggles and visors: See "personal protective equipment – general principles". |
| *Devices* | Devices that are taken out of the room:<br><br>Bladder scanner, ECG-machine, scale etc. should be cleaned and disinfected before leaving the isolation room, whenever possible. |
| *Equipment and bed linen in cupboards* | Correct hand hygiene should be performed (i.e. change of gloves and hand hygiene) before handling equipment and linen in cupboards.<br><br>If equipment and bed linen in cupboards have not been handled correctly, cf. above, it should be send for retreatment or wash at the laundry. Disposable items are discarded. |
| *Cleaning after procedures* | Spill and visible contamination by organic material (e.g. faeces/vomit): Spill is wiped up, followed by cleaning and disinfection.<br><br>Procedures with high risk of aerosol generation from the airways of the patient: Horizontal surfaces and contact points within 2 meters' distance from the patient's airways are cleaned and disinfected: |

| | |
|---|---|
| | • For repeated procedures: 3 times a day and always at visible contamination<br>• For single procedures: After the procedure is completed |
| *Daily cleaning* | Service staff are informed of the isolation and must use PPE as the other personnel.<br><br>Room, bedside table, other furniture and equipment/devices close to the patient are cleaned.<br><br>Contact points (handles, guard rail, headboard, elevation remote control, pull cord, switches on lamps/devices/apparatus) are cleaned and disinfected.<br><br>Bath and toilet are cleaned. Contact points (handles, taps, toilet seats and toilet flush button) are cleaned and disinfected.<br><br>The floor is cleaned.<br><br>Cleaning equipment: mops and cloths are wrapped up in the room and sent for washing. Only necessary equipment and cloths/cleaning agents are brought to the room. Equipment are cleaned and disinfected before taken out of the room, alternatively they are taken straight to a washer disinfector. |
| *Length of isolation* | **Patients with suspected COVID-19:** Isolation is discontinued, when the diagnosis is rejected. See "COVID-19. Diagnosis and treatment of, regional guideline, section 3. Diagnostics COVID-19".<br><br>**Patients with a mild or moderate disease course of COVID-19:**<br><br>Isolation can be discontinued when *one* of the following criteria is fulfilled:<br><br>• End of symptoms ≥ 48 hours<br><br>OR<br><br>• From 10 days after symptom onset, provided fever-free for 48 hours (without antipyretic medication) with significant clinical improvement, and thus only mild recurring symptoms as cough, loss of sense of taste and smell, headache, fatigue etc.<br><br>**Patients with a severe disease course of COVID-19** (need for intensive care, including respirator and ultimately ECMO treatment) and immunocompromised patients: Isolation can be discontinued based on a concrete medical assessment. See "COVID-19. Diagnosis and treatment of, regional guideline, section 6. Discontinuation of isolation".<br><br>**Patients without symptoms of COVID-19 who have tested positive** for SARS-CoV-2 RNA: Isolation can be discontinued 7 days after the first positive test provided that the patient is still without symptoms of COVID-19. This is independent on possible later tests.<br>**Patients without symptoms of COVID-19 who are close contacts** to a SARS-CoV-2 positive person: The patient is tested on day 4 and 6 after last exposure and isolation can be discontinued if the test for SARS-CoV-2 RNA on day 6 is negative, provided that the patients has not developed symptoms. |
| *Final cleaning of room and furniture* | When the patient has been discharges from the room, a final cleaning is performed<br><br>Service staff are informed of the isolation and must use long-sleeve disposable gown and gloves when cleaning the room. By risk of splatter and |

| | |
|---|---|
| | droplets full face shield or face mask type II and eye protection should additionally be used.<br><br>Only necessary equipment and cloths/cleaning agents are brought to the room. Equipment is cleaned and disinfected before leaving the room.<br><br>The bed is emptied at the room:<br><br>• Duvets and pillows are sent for wash in a laundry bag.<br>• Pillows with washable surfaces are cleaned and disinfected at the room.<br>• The bed (guard rails, head- and footboards, bed frame, remote controls, possible IV pole and lifting pole) is cleaned and disinfected.<br>• Mattress including borders is cleaned and disinfected.<br>• The bed is then cleaned thoroughly.<br><br>The room is cleaned. Horizontal surfaces and contact points, bedside table, all devices/apparatus and aids are cleaned and disinfected.<br><br>Bath and toilet are cleaned. Toilet, sink, and shower fixtures are disinfected, Toilet paper and toilet brush are thrown away.<br><br>Shower curtain, bed curtain, and curtains are sent for wash in a laundry bag.<br><br>Unused equipment, that has been available during the isolation are thrown away or reuse treated. Unused equipment from cupboards are sent for reuse treatment (disposable items are thrown away) if it has been handled without use of correct hand hygiene.<br><br>The floor is cleaned.<br><br>Mops and cloths are wrapped up at the room and sent for wash. |
| *Waste and laundry* | Bin bag and laundry bag are wrapped up at the room.<br><br>Laundry is sent for washing as usual.<br><br>Waste is handled in accordance to the hospital's usual guidelines. |
| *Deceased* | When handling a deceased person, identical guidelines as for isolation apply (see section "personal protective equipment").<br><br>Bedlinen and patient clothes must be changed.<br><br>Transportation of the deceased see "Transportation – in-house". The deceased should not wear a face mask during transport. The isolation sign regarding relevant personal protective equipment and disinfection agent are put on the bed – alternatively written on the death note.<br><br>Section and handling in the chapel, a long-sleeve disposable gown and gloves are used. Procedures with risk of splatter and droplets require the use of a full face shield or face mask type II and eye protection.<br><br>Recommendations for visitors: Avoid contact with mucous membranes. Perform hand disinfection or hand wash after touching. |

From AUH e-Dok COVID-19. Infection control precautions, regional guideline. November 26, 2020

### Schema 2: Outpatient, diagnostic, and treatment units

General infection control guidelines always apply

| | |
|---|---|
| *Outpatient, diagnostic, or treatment units receiving an isolation patient with confirmed or suspected COVID-19* | Preparation:<br><br>Furniture and equipment within a 2-meter distance from the patient are removed or covered up (or cleaned and disinfected after examination/treatment).<br><br>For choice of personal protective equipment, see appendix: "Choice of personal protective equipment by suspected or verified COVID-19<br><br>For testing units, see appendix "personal protective equipment at testing units_throat swap by suspected COVID-19<br><br>Cleaning/disinfection<br><br>• Horizontal surfaces in the room are cleaned<br>• Used apparatus and equipment (bed, chairs etc.) and contact points (places touched by the patient or personnel, e.g. handles, switches) are cleaned and disinfected.<br>• Unused equipment, that has been available during the isolation are thrown away or reuse treated. Unused equipment from cupboards are sent for reuse treatment (disposable items are thrown away) if it has been handled without use of correct hand hygiene.<br>• Spilling or visible contamination with biological material (e.g. faeces/vomit/blood/spinal fluid): Spill is wiped up. Cleaning and disinfection are done.<br><br>The floor is cleaned as needed.<br><br>Cleaning equipment: mops and cloths are wrapped up at the room and sent for washing. Only necessary equipment and cloths/cleaning agents are brought to the room. Equipment are cleaned and disinfected before taken out of the room, alternatively straight to a washer disinfector. |

From AUH e-Dok COVID-19. Infection control precautions, regional guideline. November 26, 2020

### Schema 3: Contact tracing of close contacts and outbreak management in detection of SARS-CoV-2 RNA in staff and patients

In need of counselling regarding close contacts at the hospital and suspicion of local spread of SARS-CoV-2: Contact the local infection control unit or Department of Clinical Microbiology.

| | |
|---|---|
| *When to start contact tracing of close contacts or transmission reduction* | Contact tracing of close contacts:<br>• always when a person tests positive for SARS-CoV-2 RNA, including health personnel, outpatients and inpatients.<br>Transmission reduction starts if there is a suspicion of local infection spread by SARS-CoV-2, including<br>• if an inpatient tests positive for SARS-CoV-2 RNA with reasons to believe that the patient was infected at the at the hospital<br>• if two or more patients and/or personnel test positive for SARS-CoV-2 RNA within a defined group and within a defined time period. |
| *Temporal definition of transmission reduction and contact tracing of close contacts* | Transmission reduction and contact tracing of close contacts is limited in time to 48 hours before the positive test for asymptomatic persons and 48 hours before symptom onset for symptomatic persons. |
| *Close contacts and other contacts* | For contact tracing of close contacts outside the hospital and management of close contacts in general refer to "COVID-19. Test of close contacts, regional guideline".<br>Personnel, who are close contacts are sent home* when recognized and requested to self-isolate until showing a negative test for SARS-CoV-2 RNA taken minimum 6 days (144 hours) after last exposure.<br><br>Inpatients, who are close contacts are sent isolated* until showing a negative test for SARS-CoV-2 RNA taken minimum 6 days (144 hours) after last exposure.<br><br>*Personnel and patients who are fully vaccinated or past SARS-CoV-2 infection within the last 12 months can refrain from self-isolation, if they are without symptoms of COVID-19. See "COVID-19. Test of close contacts, regional guideline".<br><br>When are healthcare personnel close contacts?<br>Persons in the health care system (personnel and patients) are considered close contacts if believed in high risk of being exposed to droplet transmission or direct/indirect contact transmission and the general infection control guidelines have not been followed – see "General infection control precautions – during an epidemic, regional direction.<br><br>If the person showing to be SARS-CoV-2 RNA positive used a type II mask correctly and general infection control guidelines for contact spread were followed in a given situation, there is by default no close contacts in the situation.<br><br>For patients:<br>Patients are regarded as close contacts, if a high risk of exposure to droplet transmission or direct/indirect contact transmission is considered, e.g. has shared toilet or been close to a person with verified COVID-19, or if caring or |

| | |
|---|---|
| | treatment tasks at the room have violated the general infection control guidelines. Patients sharing the same room will by default be defined as close contacts.<br><br>Other contacts:<br>There may be situations, where the criteria for being close contact is not fulfilled, but the overall evaluation suggests a risk of other contacts to the test-positive person being exposed to droplet spread or direct/indirect contact spread.<br>Examples of other contacts can be shared offices, meetings, or teaching, where an infected person has participated, and there is doubt whether guidelines for distance, hand hygiene, and cleaning has been followed.<br>These contacts are tested on the 4th and 6th day after exposure, but should not self-isolate. |
| *Transmission reduction and outbreak management* | Transmission reduction is initiated when there is a suspicion of local spread of infection with SARS-CoV-2, either if a patient is believed infected at the hospital or if two or more patients and/or staff members are tested positive for SARS-CoV-2 RNA within a defined group of people and a defined time period.<br><br>Transmission reduction involves immediate testing for SARS-CoV-2 RNA of all admitted patients at the relevant unit, as well as treatment and caring staff working at the relevant unit within 48 hours before the index person's symptom onset or first positive test. Tests are done at day 0 and day 7 after the risk of local infection spread is known.<br><br>If the index person is a patient, who within the last 48 hours has been transferred from another unit or admitted from an institution, a corresponding transmission reduction should be initiated at the other unit/institution.<br><br>If more cases are found during the transmission reduction:<br><br>• Outbreak management can be expanded to involve personnel, who have had short-term functions at the unit or with patients from the unit, e.g. supervising physicians, lab technicians, radiographs, anaesthesia staff, technical staff etc., as far as they can be identified.<br>• Contact tracing of close contacts to additional COVID-19 cases is initiated.<br><br>SARS-CoV-2 RNA testing of affected personnel and inpatients is repeated every 7th day as long as additional SARS-CoV-2 RNA positive persons are identified at the unit. There can be a need for a more frequent screening (e.g. 2 times per week).<br><br>While transmission reduction is ongoing, it is recommended as a supplement to also test patients who are transferred to other units/departments. Transfer should not await the answer of the SARS-CoV-2 RNA test. |

From AUH e-Dok COVID-19. Infection control precautions, regional guideline. November 26, 2020

### Appendix: Choice of personal protective equipment for suspected or verified COVID-19

The scheme solely states the **supplementing** infection control guidelines for COVID-19

**Always perform a risk assessment:**
The work task is evaluated according to duration and exposure for droplet and aerosol generating procedures. If the risk for moist or contamination is high, a fluid resistant lab coat and a full face shield combined with the face mask should be chosen. Examples can be work tasks in close proximity to a heavy coughing or sweating patient or aerosol generating procedures such as frequent inhalations.

| **Situation**<br><br>Distance from the patient's head | **Gloves and lab coat with long sleeves**<br><br>Lab coat: fluid resistant or fluid proof lab coat or apron with long sleeves. | **Type II face mask and eye protection**<br><br>Eye protection: visor, googles, or full face shield | **Type I face mask for the patient** | **FFP3/FFP2 respirator and eye protection**<br><br>Eye protection: visor, googles, or full face shield |
|---|---|---|---|---|
| > 2 m from the patient with no contact with equipment, surfaces etc. (e.g. conversation or visitation/evaluation) | No | No | No | No |
| > 2 m from the patient but with contact to equipment, surfaces etc. | Yes | No | No | No |
| < 2 m from the patient, care-taking, examination- or treatment tasks | Yes | Yes | No | No |
| < 2 m from the patient with respiratory procedures with low-risk of aerosol generation* (throat swab, medication by nebulizer, low-flow nasal oxygen/atmospheric air, Lomholt humidifier, lung physiotherapy incl. PEP-flute, lung function test, examination and treatment of dysphagic patients) | Yes | Yes | No | No |
| < 2 m from the patient with respiratory procedures with high-risk of aerosol generation (intubation and extubation, manual ventilation, laryngeal mask, short-term ventilator weaning, cardiopulmonary resuscitation (Note: situations with use of only cardiac massage and defibrillator is not included), CPAP, NIV, BiPAP, tracheal suction (open system), high-flow oxygen therapy (flow rate 30-60 l/min), high frequency oscillatory ventilation (HFOV), tracheostomy and tracheostomy procedures, induced sputum and bronchoscopy, surgery and post-mortem procedures in airways with use of high-speed rotating equipment) | Yes | No | No | Yes |
| Transportation without direct contact, the patient wearing a mask | No | No | Yes | No |
| Transportation with direct patient contact | Yes | Yes | Yes<br>If possible, to minimize con-tamination to surroundings | No |
| * Stay within 2 meters should be minimized with cough inducing procedures | | | | |

From AUH e-Dok COVID-19. Infection control precautions, regional guideline. December 4, 2020

**Table S1.** Incidence rate ratios of SARS-CoV-2 infection by population characteristics

| Characteristic | Person-days | Positive SARS-CoV-2 PCR tests | Infection rate per 100 000 person-days | Incidence rate ratio (95% CI)* |
|---|---|---|---|---|
| Sex | | | | |
| Women | 458 636 | 140 | 3.1 | 0.89 (0.55 to 1.44) |
| Men | 55 529 | 19 | 3.4 | Reference |
| Age (per year) | 514 165 | 159 | 3.1 | 0.99 (0.98 to 1.00) |
| Geographical area | | | | |
| West | 87 223 | 30 | 3.4 | 1.27 (0.74 to 2.18) |
| Central | 55 694 | 22 | 4.0 | 1.46 (0.82 to 2.61) |
| East | 282 321 | 83 | 2.9 | 1.09 (0.69 to 1.71) |
| General | 88 779 | 24 | 2.7 | Reference |
| Month | | | | |
| November | 16 208 | 8 | 4.9 | Reference |
| December | 138 941 | 73 | 5.3 | 1.06 (0.51 to 2.21) |
| January | 138 446 | 61 | 4.4 | 0.89 (0.43 to 1.87) |
| February | 88 395 | 9 | 1.0 | 0.21 (0.08 to 0.53) |
| March | 73 920 | 5 | 0.7 | 0.14 (0.04 to 0.42) |
| April | 58 255 | 3 | 0.5 | 0.10 (0.03 to 0.39) |
| PCR tests 1 to 2 days earlier | | | | |
| 0 | 442 978 | 141 | 3.2 | Reference |
| 1 | 70 916 | 18 | 2.5 | 0.80 (0.49 to 1.30) |
| 2 | 271 | 0 | - | - |
| PCR tests 3 to 7 days earlier | | | | |
| 0 | 349 800 | 91 | 2.6 | Reference |
| 1 | 156 388 | 58 | 3.7 | 1.43 (1.03 to 1.98) |
| ≥ 2 | 7977 | 10 | 12.5 | 4.82 (2.51 to 9.26) |
| PCR tests ≥ 8 days earlier | | | | |
| 0 | 54 991 | 14 | 2.5 | Reference |
| 1- 4 | 227 196 | 83 | 3.7 | 1.43 (0.81 to 2.53) |
| 5 – 9 | 151 419 | 53 | 3.5 | 1.37 (0.76 to 2.48) |
| ≥10 | 80 559 | 9 | 1.1 | 0.44 (0.19 to 1.01) |
| Department | | | | |
| Emergency | 11 922 | 4 | 3.4 | 1.38 (0.48 to 4.00) |
| Medicine | 130 099 | 54 | 4.2 | 1.71 (1.05 to 2.79) |

| | | | | |
|---|---|---|---|---|
| Surgery | 89 439 | 24 | 2.7 | 1.11 (0.62 to 1.96) |
| Biochemistry | 42 232 | 15 | 3.6 | 1.46 (0.76 to 2.81) |
| Service | 7160 | 3 | 4.2 | 1.73 (0.52 to 5.75) |
| Anaesthesiology | 19 446 | 5 | 2.6 | 1.06 (0.40 to 2.79) |
| Radiology and Nuclear Medicine | 23 074 | 8 | 3.5 | 1.43 (0.64 to 3.19) |
| Psychiatry | 62 987 | 15 | 2.4 | 0.98 (0.51 to 1.88) |
| Departments with less frequent patient contact | 94 785 | 23 | 2.4 | Reference |
| Other | 32 873 | 8 | 2.4 | 1.00 (0.45 to 2.24) |
| Occupation | | | | |
| Nursing staff | 194 191 | 78 | 4.0 | 1.94 (1.08 to 3.49) |
| Medical doctors | 46 666 | 17 | 3.6 | 1.76 (0.85 to 3.62) |
| Biomedical Laboratory | 40 055 | 12 | 3.0 | 1.45 (0.66 to 3.17) |
| Medical secretaries | 62 718 | 13 | 2.1 | Reference |
| Other | 170 387 | 39 | 2.3 | 1.10 (0.59 to 2.07) |
| Smoking | | | | |
| Current smoker | 28 449 | 8 | 2.8 | 0.99 (0.48 to 2.04) |
| Previous smoker | 150 071 | 55 | 3.7 | 1.29 ( 0.93 to 1.80) |
| Never smoker | 331 795 | 94 | 2.8 | Reference |
| BMI (kg/m2) | | | | |
| <20 | 34 027 | 8 | 2.4 | 0.78 (0.38 to 1.63) |
| 20 to 24 | 240 220 | 72 | 3.0 | Reference |
| 25 to 29 | 154 013 | 48 | 3.1 | 1.04 (0.72 to 1.50) |
| ≥ 30 | 81 850 | 29 | 3.5 | 1.18 (0.77 to 1.82) |
| Hay fever | | | | |
| No | 409 242 | 130 | 3.2 | Reference |
| Yes | 104 923 | 29 | 2.8 | 0.87 (0.58 to 1.30) |
| Asthma | | | | |
| No | 477 838 | 145 | 3.0 | Reference |
| Yes | 36 327 | 14 | 3.9 | 1.27 (0.73 to 2.20) |
| COPD | | | | |
| No | 510 886 | 158 | 3.1 | Reference |
| Yes | 3279 | 1 | 3.0 | 0.99 (0.14 to 7.04) |
| *Crude model | | | | |

**Table S2.** Day-by-day non-compliance (%) with personal protective guidelines by procedures

| Consultations (n= 63 263) | Procedures with physical contact (n=68 313) | Respiratory procedures (n=10 552) | Surgical procedures or deliveries (n=8753) | Transportation of patients (n=3601) | Other tasks within 2 meters from patients (N=20 401) | Cleaning of patient rooms (n=12 530) | Total (N=187 413) |
|---|---|---|---|---|---|---|---|
| 1.6 | 2.3 | 4.8 | 1.9 | 0.7 | 1.7 | 0.6 | 2.0 |

**Table S3.** Close contact 3-7 days earlier with patients, co-workers and persons outside work with COVID-19 and incidence rate ratios of SARS-CoV-2 infection including only COVID-19 contact information obtained before PCR test results were available

| Contact with person with COVID-19 | Person-days* | Positive SARS-CoV-2 PCR tests | Infection rates per 100 000 person-days | Incidence rate ratio (95% CI) | | | |
|---|---|---|---|---|---|---|---|
| | | | | Model 1† | Model 2‡ | Model 3§ | Model 4¶ |
| Patients | | | | | | | |
| No contact | 489 009 | 120 | 24.5 | Reference | Reference | Reference | Reference |
| Contact | 25 164 | 39 | 155.0 | 6.32 (4.40 to 9.06) | 4.65 (3.22 to 6.70) | 4.23 (2.93 to 6.12) | 3.52 (2.41 - 5.13) |
| Persons outside work | | | | | | | |
| No contact | 512 069 | 144 | 28.1 | Reference | Reference | Reference | Reference |
| Contact | 2104 | 15 | 712.9 | 25.35 (14.90 to 43.15) | 18.31 (10.70 to 31.33) | 15.11 (8.80 to 25.96) | 14.19 (8.27 - 24.33) |

*The total number of person-days is increased by 8 compared with table 3. This represents the net effect of i. an increase in person-days with missing information on contact with patients and persons outside work with COVID-19 because of the restriction in contact information and ii. a reduction in person-days with missing information because contact with co-workers with COVID-19 was not included in the models.
†Crude model
‡ Adjusted for age (continuous), sex and month (6 categories, November 2020-April 2021)
§As model 2 and additionally adjusted for the other types of COVID-19 contact
¶ As model 3 and additionally adjusted for number of previous PCR tests
